# What effect might border screening have on preventing importation of COVID-19 compared with other infections? A modelling study

**DOI:** 10.1101/2020.07.10.20150664

**Authors:** Declan Bays, Emma Bennett, Thomas Finnie

## Abstract

The effectiveness of screening travellers during times of international disease outbreak is contentious, especially as the reduction in the risk of disease importation can be very small. Border screening typically consists of travellers being thermally scanned for signs of fever and/or completing a survey declaring any possible symptoms prior to admission to their destination country; while more thorough testing typically exists, these would generally prove more disruptive to deploy. In this paper, we describe a simple Monte Carlo based model that incorporates the epidemiology of COVID-19 to investigate the potential benefit of requiring all travellers to undergo thorough screening upon arrival. This is a purely theoretical study to investigate whether a single test at point of entry might ever prove to be a way of significantly decreasing risk of importation. We therefore assume ideal conditions such as 100% compliance among travellers and the use of a “perfect” test. In addition to COVID-19, we also apply the presented model to simulated outbreaks of Influenza, SARS and Ebola for comparison. Our model only considers screening implemented at airports, being the predominant method of international travel. Primary results showed that in the best-case scenario, screening may expect to detect 8.8% of travellers infected with COVID-19, compared to 34.8.%, 9.7% and 3.0% for travellers infected with influenza, SARS and Ebola respectively. While results appear to indicate that screening is more effective at preventing disease ingress when the disease in question has a shorter average incubation period, our results indicate that screening alone does not represent a sufficient method to adequately protect a nation from the importation of COVID-19 cases.

**Data availability:** All results described in the work, in addition to technical descriptions of methods used, are made available in the supplementary material. The Python package used to implement these methods and obtain our results has been made accessible online[1].

## Introduction

While international trading and tourism has huge sociological and economic benefits, it also markedly increases the vulnerability of national populations to emerging and re-emerging infectious diseases. In particular, the ability to travel between almost any two points on the planet within 24 hours provides the potential for epidemics to rapidly evolve into pandemics[2]–[4]. On the 31^st^ December 2019, the Wuhan Municipal Health Commission reported the first cluster of individuals infected with COVID-19, at the time being an unknown pneumonia inducing disease[5]. By the end of January 2020, cases of COVID-19 had been reported in 26 countries outside of China[6]. Less than 6 weeks later, with cases being reported in 114 countries and territories, the World Health Organisation declared the COVID-19 outbreak a pandemic[7].

The World Health Organisation (WHO) recommends in its International Health Regulations[8] that all WHO States should have the capability to implement some form of screening at international points of entry during times of outbreak. Such screening has previously involved using thermal cameras to scan for signs of fever and asking travellers to self-declare any signs of symptoms. These methods are both less than perfect and have led many to disagree with the idea that border screening is a worthwhile endeavour. In this paper, we look to use mathematical modelling to evaluate whether the implementation of a “perfect” border screening policy could theoretically provide any protection from disease ingress during the ongoing outbreak. We present a simple mechanistic model that represents the process of a COVID-19 infected traveller attempting to undertake international travel and gain entry to some destination country where such a border screening policy is being enforced. The model is then run repeatedly utilising Monte Carlo simulation, capturing the stochastic nature of the various processes involved, to calculate the likelihood that an infected person would be presenting detectable symptoms upon arrival at the border of the destination country. The model we produce is easily extendable to other diseases and as such, we apply our model to simulated outbreaks of Influenza, SARS and Ebola for comparison.

## Assumptions

Work presented in the following is based upon the subsequent set of assumptions:

- All simulated individuals are assumed to have been infected prior to travelling,
- The distribution of time of infection, *D*_exp_, is uniform across the ranges 0-72, 0-168 and 0- 336 hours prior to flying, simulating where infection has occurred during a short break, a holiday, or more longer-term travel
- Border screening only detects travellers following a period of incubation; prior to completion of this they are not detectable
- Exit screening is being enforced in the country of origin so persons who have become detectable before boarding their flight do not fly
- All persons travelling only take directs flight from their country of origin to the destination country
- The distribution of flight times, *D*_flight_, are uniform across the ranges 3-5, 7-9 and 11-13 hours to represent short, medium and long-haul flights respectively.
- All people attempting to cross the border are screened
- Screening does not produce false negatives
- The number of infected persons remains constant throughout the simulation (transmission and death are neglected)
- Screening detects all infected persons who have incubated
- Infected people do not attempt to “game” the system by concealing signs of infection

We reiterate that the presented model assumes a “perfect” border screening process is being used, so the above assumptions have been chosen to reflect this.

## Methods

Our model uses Monte Carlo methods to approximate the likelihood that infected travellers, attempting to travel from country A to country B, would be detected on arrival to country B following a range of infection and travelling scenarios. We simulate a large number of infected travellers, each of which is assigned a time of infection (*t*_exp_), an incubation period (*t*_inc_) and a flight time (*t*_flight_), sampled from the distributions *D*_exp_, *D*_inc_ and *D*_flight_ respectively. These distributions are given as parameters to the model and represent the scenario being considered (note that the disease being considered is characterised in the model solely through the incubation period distribution provided here). For each traveller, these values are compared to determine whether they would become detectable prior to departure, during transit or post arrival. Results are then compiled, disregarding the travellers that would be detectable prior to departure, to determine what probability that infected travellers would detectable (and thus detected) by a perfect screening process deployed at country B’s border given that they manage to board their flight. More explicitly:

- If *t*_inc_ < *t*_exp_, the traveller has become detectable before boarding their flight and therefore does not travel (either by not being well enough to fly, or being picked up at exit screening); they exit the model being recorded as a non-flier
- If *i*_exp_ < *t*_inc_ < *t*_exp_ + t_flight_, the traveller has become detectable in transit and will therefore be detected by screening at country B’s border; they exit the model being recorded as a border-detection
- Else *t*_inc_ > *t*_exp_ + *t*_flight_, and the traveller has not become detectable prior to arriving at country B’s border and thus crosses into country B undetected; they exit the model being recorded as undetected.

This is visualised in figure 1:

**Figure 1:**
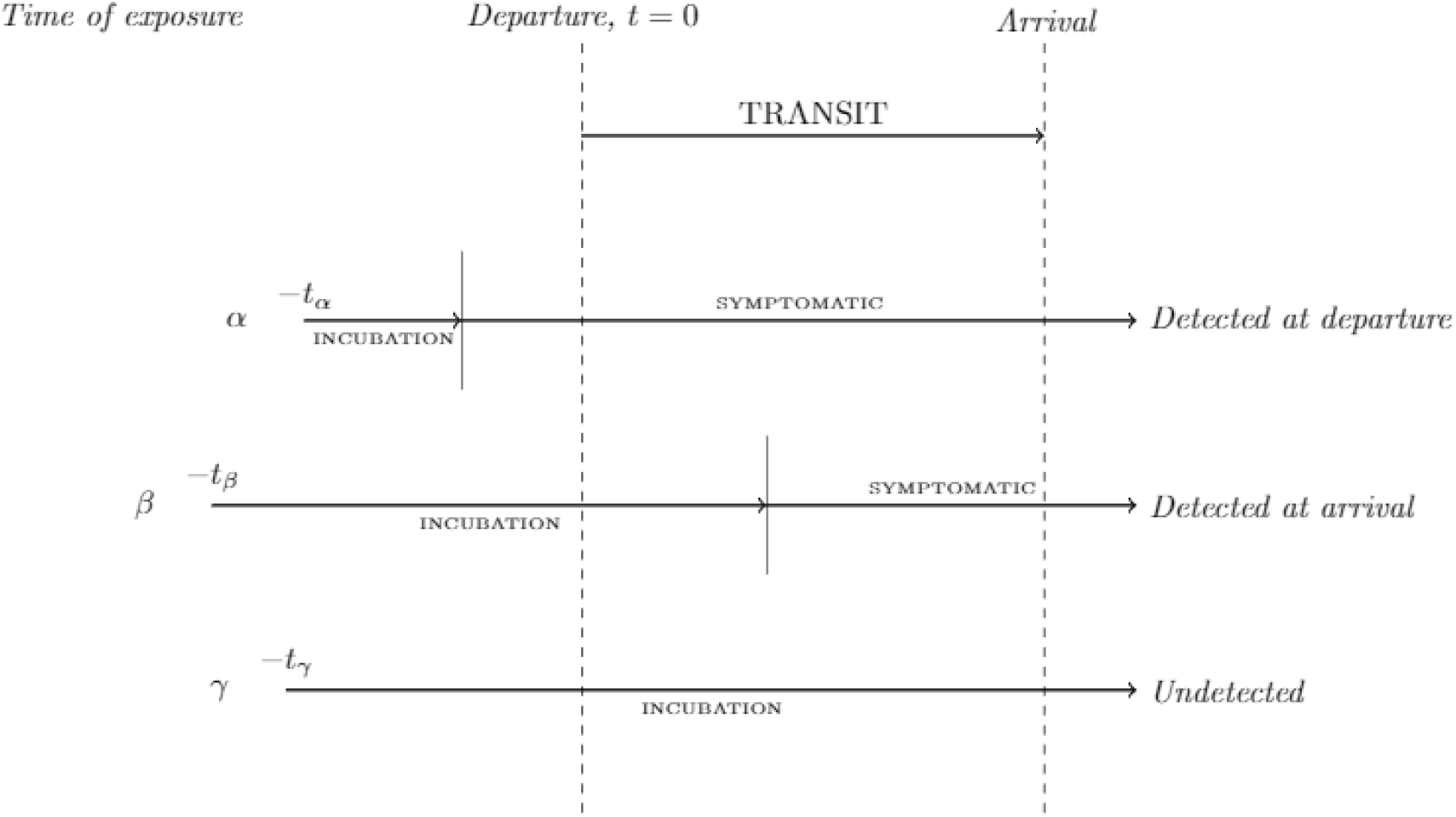
Depiction of the evaluation of individuals in the border screening model.

Each scenario is evaluated by simulating 1,000,000 infected individuals. We then take the ratio of border-detections against number of infected persons who manage to board their flight to get an approximate probability that border screening will capture infected travellers. A pseudo-code breakdown of this algorithm is included in the supplementary text, while the Python package used to implementing the above model has been made openly available online[1].

### Screening for COVID-19

The below results were obtained by applying our model to COVID-19 across all combinations of travel and infection scenarios described above. The incubation period distribution has been modelled using a log-normal distribution with parameters µ=1.6112 and σ=0.47238, which was obtained by parameterising results taken from [9] (method of parameterisation is included in the supplementary text)

**Table 1:**
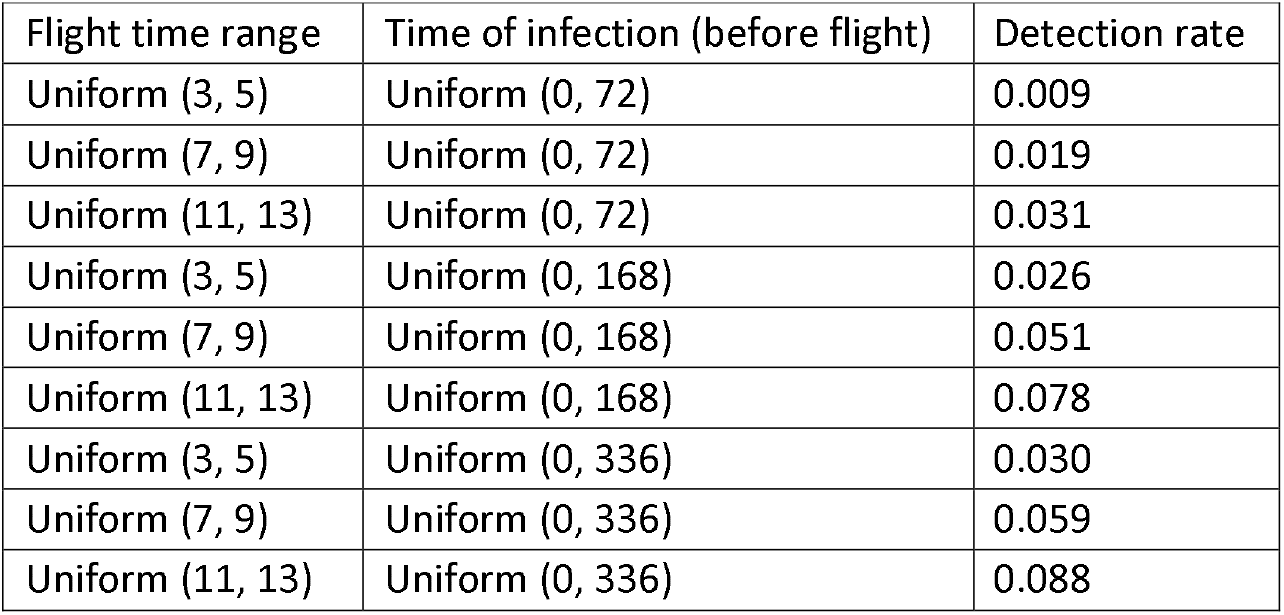
Detection rates for COVID-19 across considered scenarios.

### Screening for Influenza, SARS and Ebola

We repeat the above for simulated outbreaks of Influenza, SARS and Ebola, while also including results for COVID-19. As the rest of the method remains applicable, we need only substitute in incubation period distributions for each of these diseases. These have been taken from [10], [11] and [12] for Influenza, SARS and Ebola respectively (for derivation of gamma distribution parameters see supplementary text). For brevity, we have averaged the results across flight time ranges for each disease (table containing full set of results is included in supplementary text)

**Table 2:** Detection rates for Influenza, SARS, COVID-19 and Ebola across considered scenarios.

|  | Flight time range | Time of infection (before flight) |  |  |
| --- | --- | --- | --- | --- |
|  |  | Uniform (0, 72) | Uniform (0, 168) | Uniform (0, 336) |
| Influenza | Uniform (3, 5) | 0.116 | 0.117 | 0.116 |
|  | Uniform (7, 9) | 0.232 | 0.234 | 0.232 |
|  | Uniform (11, 13) | 0.345 | 0.346 | 0.348 |
| SARS | Uniform (3, 5) | 0.010 | 0.028 | 0.032 |
|  | Uniform (7, 9) | 0.022 | 0.056 | 0.064 |
|  | Uniform (11, 13) | 0.035 | 0.085 | 0.097 |
| COVID-19 | Uniform (3, 5) | 0.009 | 0.026 | 0.030 |
|  | Uniform (7, 9) | 0.019 | 0.051 | 0.059 |
|  | Uniform (11, 13) | 0.031 | 0.078 | 0.088 |
| Ebola | Uniform (3, 5) | 0.000 | 0.002 | 0.010 |
|  | Uniform (7, 9) | 0.000 | 0.004 | 0.020 |
|  | Uniform (11, 13) | 0.000 | 0.007 | 0.030 |

## Discussion

With our best considered scenario suggesting that screening for COVID-19 would detect less than 9% of infected travellers, it is quite plain that our model seems to indicate that the implementation of a single screening process is not sufficient to cause a significant reduction in the expected number of infected travellers entering a destination country during the COVID-19 pandemic. Detection rates also clearly decrease with average flight time, meaning screening would be even less effective on travellers arriving via short haul flights (which would presumably be the most numerous). The intuitive reason for such minimal detection rates would be, considering the average incubation time of COVID-19, that the amount of extra time afforded to individuals by their flight is not substantial enough to expect a notable proportion of infected travellers to complete their incubation period and become detectable prior to arrival (hence also the decrease with shorter flight times).

**Figure 2:**
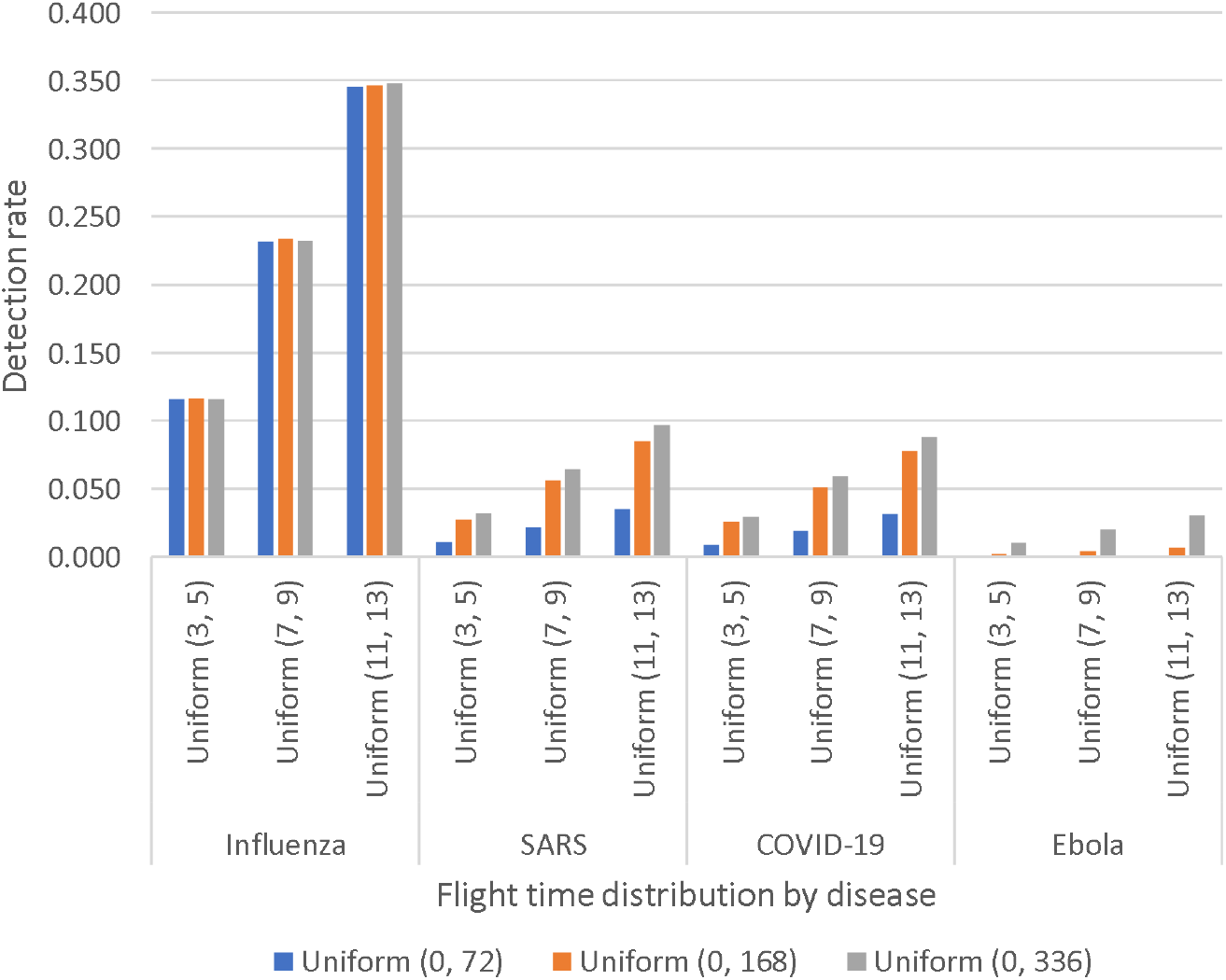
Modelled detection rates for each of the considered disease.

This argument also appears to be supported by the results obtained when applying our model to outbreaks of Influenza, SARS and Ebola; detection rates for which are plotted in the above graph. Comparing detection rates between COVID-19 and SARS first (being related diseases that have similar incubation periods), we see that detection rates are roughly in the same ballpark across all scenarios, with screening still detecting less than 10% in the best case. However, when we consider COVID-19 against Influenza or Ebola (both have markedly shorter and longer average incubation period respectively), we see from the above that in the best case we may expect to detect just under 35% of influenza cases and 3% of Ebola cases. What this could indicate is that border screening might present a viable intervention for diseases that have incredibly fast incubation periods (such as on the scale of hours), or that incorporating some additional step that provides travellers with additional time in which they might incubate (such as isolating on arrival) might make this a more successful undertaking. However, a reduction of at most 9% of arriving COVID-19 cases would be a hard sell to any public health team considering the potential cost. We would therefore conclude by stating that the results presented imply that border screening, as we have described it in this paper, would not serve as suitable intervention to prevent the ingress of further COVID-19 cases. Furthermore, while a possible 35% detection rate for influenza might seem sizeable in comparison to the results from the COVID-19 modelling, it still does not offer a reduction on the scale that might be anticipated to fully safeguard a nation from the threat of an external outbreak, with a similar conclusion being reached on all of the other of the diseases considered.

The model we described and used to obtain these results is technically simple and can therefore be rapidly evaluated with modern computation. Additionally, this simplicity allows our model to remain flexible, and easily amended to consider disease, infection and travelling scenarios outside of those considered in this work. One disadvantage though, although such considerations lay outside the remit of this work, is that the model does not consider the effects of personal behaviours (i.e. infected persons attempting to obscure signs of their infection during screening) or disease dynamics (i.e. the infection of fellow travellers during transit). However, our work sought to provide an upper bound to the potential benefit of border screening, and as such considerations would act to only decrease expected detection rates, these neglections are appropriate for the aims of this work. For the consideration of more realistic scenarios, these factors could in future be implemented into an extended version of this model.

## Conclusion

In this paper, we have presented a simple and adaptable Monte Carlo based model which can be rapidly evaluated across a range of outbreak scenarios. We then used this model to assess whether border screening could in theory provide nations with any notable protection from international travellers infected with COVID-19. Our model assumed the implementation of a perfect screening process and was applied across a range of infection and travel scenarios. Despite this, our model indicated that nations could not expect border screening alone to detect more than 9% of arriving travellers infected with COVID-19. In addition to this, we also applied the presented model to simulated outbreaks of Influenza, SARS and Ebola. Through this, while we managed to infer that screening fared slightly better when considering diseases with shorter incubation periods, results indicated that screening alone still did not offer sufficient protection from international outbreaks. This model may be in future developed by incorporating some aspect of disease transmission and/or impropriety in infected travellers.

## Data Availability

All data referred to in paper is included in presented tables

https://github.com/publichealthengland/SIRA

## Notes

### Competing Interest Statement

The authors have declared no competing interest.

### Funding Statement

Not applicable

## References

[1] “publichealthengland/SIRA.” [Online]. Available: https://github.com/publichealthengland/SIRA. [Accessed: 21-May-2021].

[2] M. Fricker and R. Steffen, “Travel and public health,” Journal of Infection and Public Health, vol. 1, no. 2. Elsevier, pp. 72–77, 01-Jan-2008, doi: 10.1016/j.jiph.2008.10.005.

[3] A. J. Tatem, D. J. Rogers, and S. I. Hay, “Global Transport Networks and Infectious Disease Spread,” Advances in Parasitology, vol. 62. pp. 293–343, 2006, doi: 10.1016/S0065-308X(05)62009-X.

[4] A. J. Tatem, “Editor’s choice: Mapping population and pathogen movements,” Int. Health, vol. 6, no. 1, p. 5, 2014, doi: 10.1093/INTHEALTH/IHU006.

[5] “Timeline of ECDC’s reponse to COVID-19.” [Online]. Available: https://www.ecdc.europa.eu/en/covid-19/timeline-ecdc-response. [Accessed: 03-Dec-2020].

[6] “Timeline of the COVID-19 pandemic in January 2020 - Wikipedia.” [Online]. Available: https://en.wikipedia.org/wiki/Timeline_of_the_COVID-19_pandemic_in_January_2020. [Accessed: 03-Dec-2020].

[7] “Situation Report-51 SITUATION IN NUMBERS total and new cases in last 24 hours.”

[8] WHO, “International Health Regulations - Third Edition,” Who, vol. 2005, p. 84, 2016, doi: 10.1017/CBO9781107415324.004.

[9] N. M. Linton et al “Incubation Period and Other Epidemiological Characteristics of 2019 Novel Coronavirus Infections with Right Truncation: A Statistical Analysis of Publicly Available Case Data.,” J. Clin. Med., vol. 9, no. 2, Feb. 2020, doi: 10.3390/jcm9020538.

[10] H. Nishiura, N. Wilson, and M. G. Baker, “Quarantine for pandemic influenza control at the borders of small island nations,” BMC Infect. Dis., vol. 9, no. 1, p. 27, Mar. 2009, doi: 10.1186/1471-2334-9-27.

[11] A. Y. C. Kuk and S. Ma, “The estimation of SARS incubation distribution from serial interval data using a convolution likelihood,” Stat. Med., vol. 24, no. 16, pp. 2525–2537, Aug. 2005, doi: 10.1002/sim.2123.

[12] W. B. P. Pettey, M. E. Carter, D. J. A. Toth, M. H. Samore, and A. V. Gundlapalli, “Constructing Ebola transmission chains from West Africa and estimating model parameters using internet sources,” Epidemiol. Infect., vol. 145, no. 10, pp. 1993–2002, Jul. 2017, doi: 10.1017/S0950268817000760.

