## Supplementary material for "What effect might border screening have on preventing importation of COVID-19 compared with other infections? A modelling study": S1 - Text

**Border Screening Model**

For our model to run computationally, we break down the model description given in the papers main body into a form easily transferable into computer code. This takes the following form:

Non_fliers = 0

Border_detected = 0

Non_detected = 0

**For** $0 \leq i \leq n$ **do**

**Sample** $\mathbf{t}_{\text{inc}}\mathbf{,}\mathbf{t}_{\text{exp}}\mathbf{,}\mathbf{t}_{\text{flight}}$ **from** $\boldsymbol{D}_{\text{exp}}\boldsymbol{,}\boldsymbol{D}_{\text{inc}}$**,** $\boldsymbol{D}_{\text{flight}}$

**If** $\mathbf{t}_{\text{inc}} \boldsymbol{\leq} \mathbf{t}_{\text{exp}}$**, do**

No_fly_count += 1

**Else if** $\mathbf{t}_{\text{inc}}\boldsymbol{\leq}\mathbf{t}_{\text{exp}}\mathbf{+}\mathbf{t}_{\text{flight}}$**, do**

**If weighted_coin_flip(**$\boldsymbol{\alpha}$**), do**

**If weighted_coin_flip(**$\boldsymbol{\beta}$**), do**

Border_detected += 1

**Else do**

Non_detected += 1

Fliers = n – Non_fliers

Detection_ratio = Border_detected / Fliers

**Return** Detection_ratio

Where we use the function **weighted_coin_flip(**$\mathbf{x}$**)**, defined to return the value True with a probability $x$, to simulate the random process of a symptomatic case being screened and subsequently detected. Note that we do not need to consider whether non-symptomatic persons undergo screening, as they will not be detectable, and this will just add processing time.

**Calculating incubation parameters for Ebola and influenza**

In the referenced papers (Nishiura et al., 2009; Pettey et al., 2017), the authors state that a gamma distribution was used to fit incubation period in both cases. However, in the Pettey paper only the mean and the 95^th^ percentile was given, while in the Nishiura paper only the mean and the variance was given.

To obtain shape and scale parameters from these values, we used two different methods for each of the different types of stated values. For both cases we applied the following formula:

$$\text{shape}=\left( \frac{\text{mean}}{\text{standard deviation}} \right)^{2}$$

$$\text{scale}=\frac{\text{(standard }\text{deviation)}^{2}}{\text{mean}}$$

For the influenza parameters, where the mean and variance (hence standard deviation) was given in the text, this was a simple case of entering the values into the formulas. For the Ebola parameters however (where only the mean and the 95^th^ percentile was given), this was done using Python to create a range of values for the standard deviation. From here, a range of values for the shape and scale parameters were then calculated using these formulas; one for each value in the range of standard deviations. A gamma distribution was then created from each pair of shape and scale parameters using pythons SciPy package. Each of these distributions was then assessed for the value of the 95^th^ percentile, and the distribution which gave a value closest to the reported value in the paper was then selected, and the corresponding parameters recorded.

**Calculating incubation parameters for COVID-19**

In table 1 of the referenced paper (Linton et al., 2020), it is given that the mean and standard deviation for the incubation period of infected persons (obtained from 158 recordings of cases which include Wuhan resident) were 5.6 and 2.8 respectively. We can then transform these values to obtain the lognormal parameters for this distribution. Our parameterisation requires the values $\mu\text{and} \sigma$, which are obtained through the formulas (see <https://en.wikipedia.org/wiki/Log-normal_distribution>):

$$\mu=\ln\left( \frac{\left( \text{mean} \right)^{2}}{\sqrt{\left( \text{standard devaition} \right)^{2}+\left( \text{mean} \right)^{2}}} \right)$$

$$\sigma^{2}=\ln\left( 1+\frac{\left( \text{standard deviation} \right)^{2}}{\left( \text{mean} \right)^{2}} \right)$$

**Full results from Influenza, SARS and Ebola modelling**

| Disease name | Incubation distribution | Exposure time range (time before flight) | Flight time range | Calculated border screening success rate |
| --- | --- | --- | --- | --- |
| Ebola | Gamma (shape: 8.27054, scale: 1.51139) | Uniform (0, 72) | Uniform (3, 5) | 4.60E-05 |
| Ebola | Gamma (shape: 8.27054, scale: 1.51139) | Uniform (0, 72) | Uniform (7, 9) | 0.000111 |
| Ebola | Gamma (shape: 8.27054, scale: 1.51139) | Uniform (0, 72) | Uniform (11, 13) | 0.000215 |
| Ebola | Gamma (shape: 8.27054, scale: 1.51139) | Uniform (0, 168) | Uniform (3, 5) | 0.002125 |
| Ebola | Gamma (shape: 8.27054, scale: 1.51139) | Uniform (0, 168) | Uniform (7, 9) | 0.004405 |
| Ebola | Gamma (shape: 8.27054, scale: 1.51139) | Uniform (0, 168) | Uniform (11, 13) | 0.006745 |
| Ebola | Gamma (shape: 8.27054, scale: 1.51139) | Uniform (0, 336) | Uniform (3, 5) | 0.009934 |
| Ebola | Gamma (shape: 8.27054, scale: 1.51139) | Uniform (0, 336) | Uniform (7, 9) | 0.020002 |
| Ebola | Gamma (shape: 8.27054, scale: 1.51139) | Uniform (0, 336) | Uniform (11, 13) | 0.030287 |
| SARS | Weibull (shape: 2.59, scale: 5.8) | Uniform (0, 72) | Uniform (3, 5) | 0.010119 |
| SARS | Weibull (shape: 2.59, scale: 5.8) | Uniform (0, 72) | Uniform (7, 9) | 0.02239 |
| SARS | Weibull (shape: 2.59, scale: 5.8) | Uniform (0, 72) | Uniform (11, 13) | 0.034457 |
| SARS | Weibull (shape: 2.59, scale: 5.8) | Uniform (0, 168) | Uniform (3, 5) | 0.027762 |
| SARS | Weibull (shape: 2.59, scale: 5.8) | Uniform (0, 168) | Uniform (7, 9) | 0.05589 |
| SARS | Weibull (shape: 2.59, scale: 5.8) | Uniform (0, 168) | Uniform (11, 13) | 0.084148 |
| SARS | Weibull (shape: 2.59, scale: 5.8) | Uniform (0, 336) | Uniform (3, 5) | 0.032959 |
| SARS | Weibull (shape: 2.59, scale: 5.8) | Uniform (0, 336) | Uniform (7, 9) | 0.060928 |
| SARS | Weibull (shape: 2.59, scale: 5.8) | Uniform (0, 336) | Uniform (11, 13) | 0.099513 |
| Influenza | Gamma (shape: :4.7556, scale:0.3007) | Uniform (0, 72) | Uniform (3, 5) | 0.118067 |
| Influenza | Gamma (shape: :4.7556, scale:0.3007) | Uniform (0, 72) | Uniform (7, 9) | 0.228719 |
| Influenza | Gamma (shape: :4.7556, scale:0.3007) | Uniform (0, 72) | Uniform (11, 13) | 0.347704 |
| Influenza | Gamma (shape: :4.7556, scale:0.3007) | Uniform (0, 168) | Uniform (3, 5) | 0.115599 |
| Influenza | Gamma (shape: :4.7556, scale:0.3007) | Uniform (0, 168) | Uniform (7, 9) | 0.231476 |
| Influenza | Gamma (shape: :4.7556, scale:0.3007) | Uniform (0, 168) | Uniform (11, 13) | 0.348121 |
| Influenza | Gamma (shape: :4.7556, scale:0.3007) | Uniform (0, 336) | Uniform (3, 5) | 0.115613 |
| Influenza | Gamma (shape: :4.7556, scale:0.3007) | Uniform (0, 336) | Uniform (7, 9) | 0.235614 |
| Influenza | Gamma (shape: :4.7556, scale:0.3007) | Uniform (0, 336) | Uniform (11, 13) | 0.346128 |

1. Emergency Response Department, Public Health England, Porton Down, Salisbury, SP4 0JG
